# Deep Learning for Automatic Segmentation of Vestibular Schwannoma: A Retrospective Study from Multi-Centre Routine MRI

**DOI:** 10.1101/2022.08.01.22278193

**Authors:** Aaron Kujawa, Reuben Dorent, Steve Connor, Suki Thomson, Marina Ivory, Ali Vahedi, Emily Guilhem, Navodini Wijethilake, Robert Bradford, Neil Kitchen, Sotirios Bisdas, Sebastien Ourselin, Tom Vercauteren, Jonathan Shapey

## Abstract

Automatic segmentation of vestibular schwannoma (VS) from routine clinical MRI has potential to improve clinical workflow, facilitate treatment decisions, and assist patient management. Previous work demonstrated reliable automatic segmentation performance on datasets of standardised MRI images acquired for stereotactic surgery planning. However, diagnostic clinical datasets are generally more diverse and pose a larger challenge to automatic segmentation algorithms, especially when post-operative images are included. In this work, we show for the first time that automatic segmentation of VS on routine MRI datasets is also possible with high accuracy.

We acquired and publicly release a curated multi-centre routine clinical (MC-RC) dataset of 160 patients with a single sporadic VS. For each patient up to three longitudinal MRI exams with contrast-enhanced T1-weighted (ceT1w) (n=124) and T2-weighted (T2w) (n=363) images were included and the VS manually annotated. Segmentations were produced and verified in an iterative process: 1) initial segmentations by a specialized company; 2) review by one of three trained radiologists; and 3) validation by an expert team. Inter- and intra-observer reliability experiments were performed on a subset of the dataset. A state-of-the-art deep learning framework was used to train segmentation models for VS. Model performance was evaluated on a MC-RC hold-out testing set, another public VS datasets, and a partially public dataset.

The generalizability and robustness of the VS deep learning segmentation models increased significantly when trained on the MC-RC dataset. Dice similarity coefficients (DSC) achieved by our model are comparable to those achieved by trained radiologists in the inter-observer experiment. On the MC-RC testing set, median DSCs were 86.2(9.5) for ceT1w, 89.4(7.0) for T2w and 86.4(8.6) for combined ceT1w+T2w input images. On another public dataset acquired for Gamma Knife stereotactic radiosurgery our model achieved median DSCs of 95.3(2.9), 92.8(3.8), and 95.5(3.3), respectively. In contrast, models trained on the Gamma Knife dataset did not generalise well as illustrated by significant underperformance on the MC-RC routine MRI dataset, highlighting the importance of data variability in the development of robust VS segmentation models.

The MC-RC dataset and all trained deep learning models were made available online.

## 1 INTRODUCTION

Vestibular Schwannoma (VS) is a slow growing, benign tumour that develops in the internal auditory canal. It originates from an abnormal multiplication of Schwann cells within the insulating myelin sheath of the vestibulo-cochlear nerve. It typically presents with hearing loss but also frequently causes tinnitus and balance disturbance. Larger tumours may also cause headaches, cranial neuropathies, ataxia, and hydrocephalus. It is estimated that 1 in 1000 people will be diagnosed with a VS in their lifetime (Marinelli et al., 2018); however, improvements in magnetic resonance imaging (MRI) that facilitate the detection of smaller VS have led to an increased incidence of VS in recent years (Stangerup et al., 2006). Treatment options include conservative management, radiosurgery, radiotherapy, and microsurgery for tumours that are growing or exhibit mass effect (Carlson et al., 2015).

Previous studies have demonstrated that a volumetric measurement is more accurate than linear measurements and smaller interval changes in VS size may be detected (Varughese et al., 2012; MacKeith et al., 2018). Implementing routine volumetric measurements would enable clinicians to more reliably demonstrate tumour growth and potentially offer earlier interventions. However, available tools make calculating tumour volume assessment a labour-intensive process, prone to variability and subjectivity. Consequently, volumetric methods of measuring tumour size have not been widely implemented in routine clinical practice (MacKeith et al., 2018).

To reduce the workload for clinical staff and free resources, deep learning models have recently been developed to automate this time-consuming and repetitive task. Shapey et al. (2019) and Wang et al. (2019) previously presented a deep learning framework for automatic segmentation of VS that achieved high accuracy on a large publicly available dataset of MR images acquired for Gamma Knife (GK) stereotactic radiosurgery. According to Shapey et al. (2019), “the main limitation of [their] study is […] that it was developed using a uniform dataset and consequently may not immediately perform as well on images obtained with different scan parameters.”

Such scan parameters include the type of pulse sequence and hardware-specific parameters relating to the MRI scanner and radio-frequency coil such as the magnetic field strength and field inhomogeneities as well as the use and type of contrast agent. These parameters influence the degree of T1-weighted (T1w) and T2-weighted (T2w) contrast, determine the image resolution and field-of-view (FOV) and regulate the image noise and other acquisition artefacts. As there are no official national guidelines for MRI acquisition protocols for VS in the UK imaging centres choose and optimize pulse sequences independently from each other so that images from different centres are rarely equivalent with scan parameters varying widely.

Although deep-learning models have pushed performance in medical image segmentation to new heights, they are particularly sensitive to shifts between the training and testing data (Van Opbroek et al., 2014; Donahue et al., 2014). Thus, a model that is trained on data from a single scanner with a fixed set of settings might perform well on images acquired with the same scanner/settings but fail on images acquired differently. Nevertheless, 3D-segmentation models for VS published to date rely on standardized radiosurgery treatment planning data from individual institutions with minimal differences in acquisition parameters (Shapey et al., 2019; Wang et al., 2019; Shapey et al., 2021c; Lee et al., 2021; Dorent et al., 2023). Previous studies have focused on retrospectively collected radiosurgery datasets because they often contain high quality verified manual segmentations required for treatment planning and dose calculation. However, standardized acquisition protocols mean that these datasets lack variability in terms of their acquisition parameters.

Furthermore, tumour characteristics in such datasets are biased towards tumours which are suitable for radiosurgery whereas tumours suitable for conservative management or microsurgery are under-represented. Post-operative cases are also typically not included in radiosurgery datasets although they account for a significant fraction of VS images in routine clinical practice as patients undergo regular follow-up scans to monitor tumour residuals and recurrence. After surgery, a disrupted anatomy in the cerebellopontine angle (CPA), accumulation of cerebrospinal fluid in the former tumour cavity, and the usually small size of residual tumour tissue can make the segmentation of post-operative VS particularly challenging. Moreover, these structural alterations vary depending on the surgical approach and the size of the resected volume which introduces significant variability in post-operative VS presentation on medical images. Consequently, deep learning models trained on standardized pre-operative datasets are unlikely to perform robustly in a general clinical setting.

In this work, we present for the first time deep learning models for automatic 3D-segmentation that perform well on routine clinical scans acquired for diagnosis and surveillance and which generalize to a wide range of scan parameters. We acquired a large multi-centre routine clinical (MC-RC) longitudinal dataset with images from 10 medical centres and devised a multi-stage, iterative annotation pipeline to generate high quality manual ground truth segmentations for all 3D images. The new dataset was used to develop segmentation models and assess their performance on a hold out subset of this MC-RC dataset and on 2 public VS datasets. Results show that the models perform robustly on most images and generalize to independent datasets. In particular, on independent datasets, they outperform earlier models trained on images that were acquired for Gamma Knife (GK) stereotactic radiosurgery when evaluated on unseen data.

In clinical practice, the models can be applied to monitor tumour size, post-operative residuals, and recurrence more accurately and efficiently, thereby facilitating the VS surveillance and management of patients. The MC-RC dataset will help to facilitate further research into automatic methods for VS diagnosis and treatment and can serve as a benchmark dataset for VS segmentation methods. The dataset and all trained deep learning models were made available online.

## 2 MATERIALS AND METHODS

### 2.1 Ethics statement

This study was approved by the NHS Health Research Authority and Research Ethics Committee (18/LO/0532). Because patients were selected retrospectively and the MR images were completely anonymised before analysis, no informed consent was required for the study.

### 2.2 Multi-Centre Routine Clinical (MC-RC) dataset

Our MC-RC dataset including all manual segmentations is available for download on The Cancer Imaging Archive (TCIA) (Kujawa et al., 2023b).

#### 2.2.1 Study population

The dataset contains longitudinal MRI scans with a unilateral sporadic VS from 10 medical sites in the United Kingdom. The data acquired at these medical sites was accessible and collected at the skull base clinic at the National Hospital of Neurology and Neurosurgery (London, UK) where all included patients were consecutively seen over an approximate period from April 2012–May 2014. All adult patients aged 18 years and above with a single unilateral VS were eligible for inclusion in the study, including patients who had previously undergone previous surgical or radiation treatment. Patients with Neurofibromatosis type 2 (NF2) were excluded. All patients had a minimum 5-year surveillance period.

#### 2.2.2 Uncurated dataset

Imaging data from 168 patients with dates of imaging ranging between February 2006 and September 2019 were screened for the study. The median number of time points at which each patient underwent an MRI examination was 4 (interquartile range (IQR) 3-7), and the median number of MRI sequences acquired per session was 7 (IQR 4-9). The complete uncurated image dataset comprised MRI sessions from 868 time points with 5805 MRI scans.

#### 2.2.3 Automatic image selection

To select the images most relevant for VS delineation and volumetry an automatic selection pipeline illustrated in Figure 1 was employed. For each patient images from at most 3 time points were included in the final dataset to limit the number of manual segmentations required. If more than 3 time points were available the first, last, and the time point closest to the midpoint were included while data from all other time points were discarded. Consequently, initial diagnostic as well as post-operative images were included in the final dataset. Images with a slice thickness of more than 3.9 mm were excluded due to the decreased sensitivity to small lesions and partial volume effects, which make accurate VS delineation and volumetric analysis difficult. Finally, for each of the remaining time points, image series were selected subject to the following selection rules which were designed to automatically select the most suitable MRI scans for manual segmentation.

**Figure 1.**
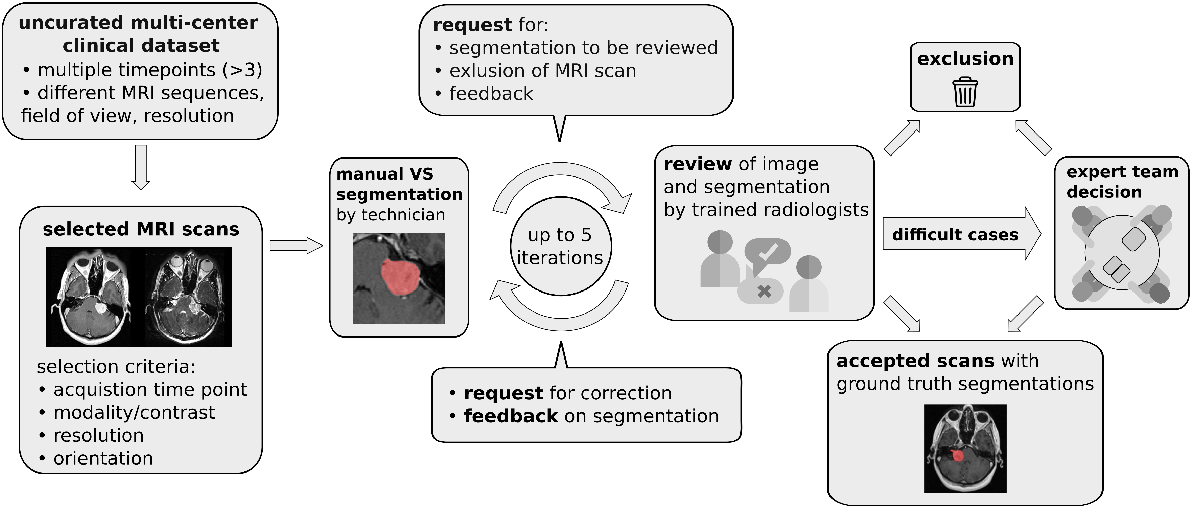
Pipeline for data curation, iterative generation of manual vestibular schwannoma ground truth segmentations, and review of the annotated multi-centre routine clinical (MC-RC) dataset.

1. If a high-resolution contrast enhanced T1 (ceT1w) image was available the image was selected. Highresolution was defined as a voxel spacing of less than 1 mm in the three directions of the voxel grid. (19 time points)
2. If a low-resolution (defined as not high-resolution) ceT1w image and a high-resolution T2w image (hrT2w) were available, both were selected. The low-resolution ceT1w image was selected with preference for axial orientation. (60 time points)
3. If a low-resolution ceT1w image was available but no hrT2w image, the low-resolution ceT1w image was selected. (45 time points)
4. If no ceT1w image was available a T2w image was selected with preference for high resolution. (303 time points)

Finally, for cases where multiple images of the same modality passed the selection process, the image with the smallest average voxel spacing was chosen.

#### 2.2.4 Exclusion of imaging data

Subsequently, during the manual annotation process, 59 time points were excluded either because parts of the tumour were outside the FOV (n=39), because a different tumour type (meningioma, trigeminal schwannoma) was identified (n=4) or because severe imaging artefacts prevented accurate VS delineation (n=4). Post-operative images in which no residual tumour could be identified (n=12) were excluded from the MC-RC training dataset, because the corresponding ground truth segmentations without foreground pixels complicate the model training and evaluation in terms of Dice Similarity Coefficient (DSC). However, model performance on these images was considered in a separate evaluation.

#### 2.2.5 Demographic data

The final MC-RC dataset after the above exclusion and curation included 160 patients (males/females 72:88; median age 58 years, IQR 49–67 years). 11 patients had imaging data from a single time point, 31 patients from two time points, and 118 patients from three time points, resulting in a total of 427 time points and 487 3D images. The average time between the first two time points was 2.4 ± 1.6 years and between the first and third time point 4.9 ± 2.7 years. With respect to the image modality, 64 time points included only a ceT1w image, 303 time points only a T2w image, and 60 time points included both. The ceT1w images comprised 19 high-resolution and 105 low-resolution images, while the T2w images comprised 349 high and 14 low-resolution images.

#### 2.2.6 Scanner/acquisition settings

Out of 427 MRI exams, 205 were acquired on a SIEMENS, 111 on a Philips, 110 on a General Electrics, and 1 on a Hitachi MRI scanner. The magnetic field strength was 1.5T for 314 exams, 3.0T for 78 exams, 1.0 T for 34 exams and 1.16T for 1 exam. Figure 2 shows the distributions of slice thickness and voxel volume and the intensity distribution of the dataset in comparison to a Gamma-Knife dataset acquired for stereotactic radiosurgery (described in Section 2.3).

**Figure 2.**
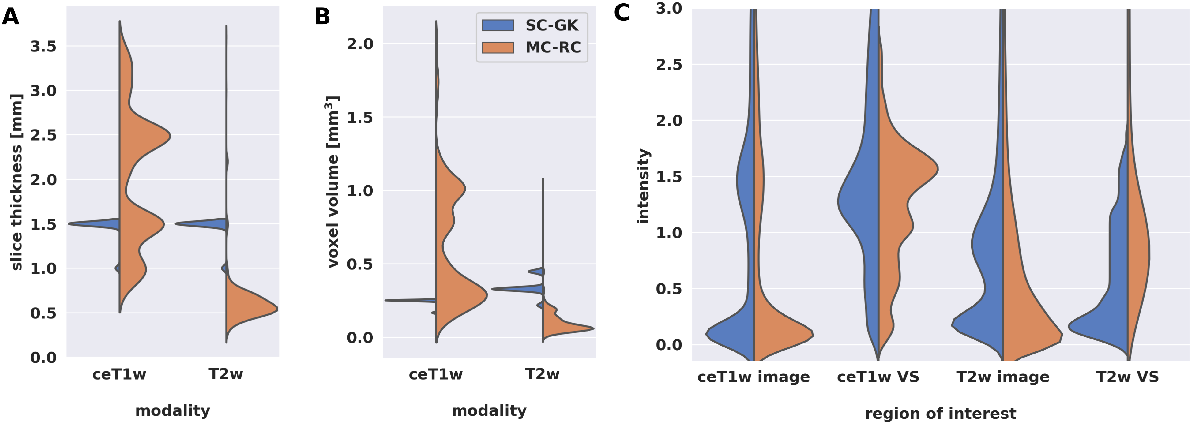
Comparison of multi-centre clinical (MC-RC) and single-centre Gamma Knife (SC-GK) datasets. Distributions of **(A)** slice thickness and **(B)** image resolution in terms of voxel volume across all ceT1w and T2w images. Parameter values of the MC-RC dataset vary significantly, while parameters of the SC-GK dataset are fixed to a small range of values. **(C)** Comparison of the normalized voxel intensity distributions of the whole image and the voxels belonging to the vestibular schwannoma (VS). The standardized acquisition protocol of the SC-GK dataset results in similar intensity distributions for each scan and hence more pronounced peaks in the average intensity distribution, while the increased variability in the MC-RC dataset leads to a wider spread of intensity values.

#### 2.2.7 Ground truth segmentations

A multi-stage manual annotation pipeline illustrated in Figure 1 was designed to obtain high quality ground truth segmentations. At the centre of the pipeline is an iterative process in which annotations were gradually improved and reviewed at each iteration. Initial VS segmentations were produced by a technician at a company specialized in providing brain measurement services based on MRI scans (Neuromorphometrics, Somerville, Massachusetts, USA) according to our specified guidelines. Focus was placed on the accuracy of segmentation edges, the brain/tumour interface, tumour within the internal acoustic meatus, the exclusion of obvious neurovascular structures from the segmentation, and for postoperative images the exclusion of scar tissue and fat. Capping cysts were included in the segmentation. If a time point included both ceT1w and T2w images, the segmentation was performed on the voxel grid of the higher resolution image and under additional visual assessment of the other image. Subsequently, each segmentation was reviewed by one of three trained radiologists (MI, AV, EM) who either accepted the segmentation or provided suggestions for improvement in the form of written comments. Alternatively, reviewers had options to exclude scans that did not fulfil inclusion criteria or refer ambiguous cases to an expert team consisting of two consultant neuroradiologists (SC + ST) and a consultant neurosurgeon (JS). During each iteration, the specialist technician improved segmentations based on the reviewer feedback until each segmentation was accepted or the corresponding image excluded from the dataset. Finally, a subset of segmentations that had either been flagged by the reviewers as ambiguous or had not been accepted after 5 iterations was reviewed and jointly annotated by the expert team. All segmentations were created, edited, and reviewed using the segmentation tool ITK-SNAP (Yushkevich et al., 2006).

#### 2.2.8 Inter- and intra-observer reliability

Inter- and intra-observer reliability was assessed on a subset of 10 ceT1w (5 high and 5 low resolution) and 41 T2w images (39 high and 2 low resolution). For intra-observer reliability assessment, 2 sets of segmentations were provided by the specialist technician at two time points, approximately 5 months apart. The first set of segmentations was reviewed according to the described iterative process, while the second set produced at the end of the learning curve for the technician was not reviewed. Thus, the measured intra-observer reliability reflects the annotators capability to recreate the first set of validated annotations.

Inter-observer reliability was based on 4 annotators. The first set of segmentations was provided by the specialist technician, while the other 3 sets were generated independently by the three reviewers (trained radiologists). For each pair of annotators and for both modalities, the mean DSC over all images in the subset was calculated and the averaged results reported.

### 2.3 Single-Centre Gamma Knife (SC-GK) dataset

This dataset was chosen as an example of a GK dataset acquired on a single scanner with little variation in sequence parameters. It enables a comparison of models trained on this dataset with models trained on our MC-RC dataset. The SC-GK dataset is a publicly available collection (Shapey et al., 2021c; Clark et al., 2013) of 484 labelled MRI image pairs (ceT1w and T2w) of 242 consecutive patients with a unilateral VS undergoing GK Stereotactic Radiosurgery. 51 patients had previously undergone surgery. Images were acquired with a 1.5T MRI scanner (Avanto Siemens Healthineers). For further details we refer to the dataset publication (Shapey et al., 2021c,b).

#### 2.3.1 Patient overlap with SC-GK dataset

While 58 patients whose MRI are included in the MC-RC dataset also have MRIs included in the SC-GK dataset, the time points and MRI series were mostly different. However, 8 series are included in both datasets and were considered separately when creating training and testing sets (Section 2.5.1). A record of overlapping patients and series is included in the MC-RC dataset (Kujawa et al., 2023b).

### 2.4 Tilburg Single-Center Gamma Knife (T-SC-GK) dataset

This dataset served as a fully independent testing set. While the previous two datasets were acquired at centres in the UK, this dataset was acquired in the Netherlands, ensuring no overlap of patients or acquisition settings/protocols. The T-SC-GK dataset was released as part of the cross-modality domain adaptation (crossMoDA) VS segmentation challenge (Dorent et al., 2023). In the present study, the challenge’s ceT1w open-access training data (n=105) (Wijethilake, 2023) was adopted as a testing set for the assessment of the ceT1w based models. The challenge’s T2w private validation data was used as a testing set for the T2w (n=32) based models. Images were acquired on a Philips Ingenia 1.5T scanner using Philips quadrature head coil. Acquisition of ceT1w images was performed with a 3D-FFE sequence with in-plane resolution of 0.8×0.8mm, in-plane matrix of 256×256, and slice thickness of 1.5 mm (TR=25ms, TE=1.82ms). Acquisition of T2w images was performed with a 3D-TSE sequence with in-plane resolution of 0.4×0.4mm, in-plane matrix of 512×512, and slice thickness of 1.0 mm (TR=2700ms, TE=160ms, ETL=50).

### 2.5 Model training and testing

#### 2.5.1 Models

Each dataset’s time points were randomly split into two subsets at a ratio of 80:20 for training and testing while assuring no patient overlap between sets. Furthermore, series overlapping between MC-RC and SC-GK datasets were placed in the respective training sets. In total, 9 deep learning models were trained and tested:

- 3 models were trained on the MC-RC training set, one for each input modality/modality set
- 3 models were trained on the SC-GK training set, one for each input modality/modality set
- 3 models were trained on a combination of both training sets (MC-RC + SC-GK) while sampling training cases from either set with equal probability, one for each input modality/modality set

The input modalities/modality sets were ceT1w, T2w, or ceT1w+T2w.

#### 2.5.2 Training

All models were trained and evaluated with nnU-Net (v2), a framework for biomedical image segmentation that yields state-of-the-art results for a wide range of public datasets used in international biomedical segmentation competitions (Isensee et al., 2021). Based on the training set, the framework automatically determines the architecture of a U-Net, a well-established type of Convolutional Neural Network (CNN) in the field of medical image segmentation (Ronneberger et al., 2015).

Models were trained with 2D and 3D U-Net configurations. A five-fold cross-validation strategy was employed, resulting in 5 sets of network weights per configuration. To this purpose the training set was split into 5 non-overlapping subsets for hyperparameter optimization. The 5 complement sets served as the network input during training. Inference was performed either with only the 3D U-Net (ensemble of 5 networks) or with an ensemble of 2D and 3D U-Nets. The best model configuration was determined by evaluating all configurations on the hyperparameter optimization sets. Model ensembling was performed by averaging the softmax outputs of all networks prior to generating the segmentation map via an argmax operation. The segmentation networks were trained for 1000 epochs where one epoch is defined as an iteration over 250 mini-batches. The mini-batch size was 2. The optimizer was stochastic gradient descent with Nesterov momentum (*μ* = 0.99). The initial learning rate of 0.01 was decayed during training according to the “poly” learning rate policy (Chen et al., 2017). The loss function was the sum of crossentropy and Dice loss (Drozdzal et al., 2016). For training scripts and the full list of hyperparameters we refer to the nnU-Net source code (https://github.com/MIC-DKFZ/nnUNet) and to the publicly available model metadata (Kujawa et al., 2023a).

#### 2.5.3 Evaluation

Each model’s performance was evaluated on all testing sets of matching modality. In total, 8 testing sets were considered:

- 3 testing sets constructed from the MC-RC dataset, one for each input modality/modality set
- 3 testing sets constructed from the SC-GK dataset, one for each input modality/modality set
- 2 testing sets constructed from the T-SC-GK dataset, one for ceT1w and one for T2w input. A testing set for combined ceT1w and T2w input was not available because the crossMoDA challenged provides unpaired images.

The sample sizes for each experiment are shown in Table 1. The trained segmentation models, example input images, and usage instructions were made available online (Kujawa et al., 2023a).

**Table 1.** Dice similarity coefficients (DSC) achieved by all models trained on the multi-centre routine clinical (MC-RC), single-centre Gamma Knife (SC-GK), and combined (MC-RC+SC-GK) training sets and evaluated on the MC-RC, SC-GK, and Tilburg single-centre Gamma Knife (T-SC-GK) testing sets. The DSC values correspond to the mean DSC over all cases in the testing sets, the errors correspond to the standard deviation. A visual representation of these results is shown in Figure 4. Moreover, the split sample sizes for training sets (including hyperparameter optimization cases) and testing sets for each experiment are shown.

| testing dataset | modality | training dataset | $n_{\text{train}}$ | $n_{\text{test}}$ | DSC[%] |
| --- | --- | --- | --- | --- | --- |
| MC-RC | ceT1w | MC-RC | 52 | | $86.9 \pm 5.9$ |
| | | SC-GK | 196 | 12 | $65.5 \pm 39.8$ |
| | | MC-RC+SC-GK | 248 | | $90.4 \pm 3.6$ |
| | T2w | MC-RC | 291 | | $85.0 \pm 15.6$ |
| | | SC-GK | 196 | 72 | $16.5 \pm 31.9$ |
| | | MC-RC+SC-GK | 248 | | $85.2 \pm 14.1$ |
| | ceT1w + T2w | MC-RC | 48 | | $75.8 \pm 27.0$ |
| | | SC-GK | 196 | 12 | $21.8 \pm 30.7$ |
| | | MC-RC+SC-GK | 248 | | $79.7 \pm 23.7$ |
| SC-GK | ceT1w | MC-RC | 52 | | $90.4 \pm 6.3$ |
| | | SC-GK | 196 | 46 | $95.2 \pm 2.2$ |
| | | MC-RC+SC-GK | 248 | | $94.8 \pm 2.1$ |
| | T2w | MC-RC | 291 | | $87.5 \pm 8.0$ |
| | | SC-GK | 196 | 46 | $92.2 \pm 3.4$ |
| | | MC-RC+SC-GK | 248 | | $92.0 \pm 3.4$ |
| | ceT1w + T2w | MC-RC | 48 | | $90.8 \pm 3.7$ |
| | | SC-GK | 196 | 46 | $95.2 \pm 2.2$ |
| | | MC-RC+SC-GK | 248 | | $95.2 \pm 2.0$ |
| T-SC-GK | ceT1w | MC-RC | 52 | | $89.9 \pm 11.6$ |
| | | SC-GK | 196 | 105 | $88.9 \pm 16.2$ |
| | | MC-RC+SC-GK | 248 | | $91.5 \pm 8.1$ |
| | T2w | MC-RC | 291 | | $85.6 \pm 8.9$ |
| | | SC-GK | 196 | 32 | $76.2 \pm 22.8$ |
| | | MC-RC+SC-GK | 248 | | $87.2 \pm 6.5$ |

##### 2.5.3.1 Evaluation metrics

The main metric applied to assess and compare the models’ segmentation performances was the commonly reported Dice similarity coefficient which is the recommended evaluation metric for semantic segmentation (Maier-Hein et al., 2022). It is defined as: 

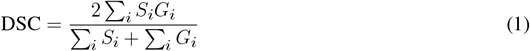

where *S*_*i*_ and *G*_*i*_ represent the binary segmentation masks of model prediction and ground truth segmentation, respectively. The DSC ranges from 0 (no overlap between model prediction and ground truth) to 1 (perfect overlap).

Additionally, we report the following metrics: average symmetric surface distance (ASSD), undirected Hausdorff distance (HD), relative absolute volume error (RVE), and distance between the centres of mass (COM).

### 2.6 Post-operative cases without residual tumour

Post-operative cases tend to be the most difficult to segment since the residual tumour is often small and obscured by scar tissue, fat, and an accumulation of CSF. In cases where no residual tumour is present the DSC is less meaningful. For example, the classification of a single voxel (or more) as a voxel belonging to the tumour would lead to a DSC of 0. Moreover, the other metrics described above are not defined for cases without residual tumour. Therefore, we examined the performance of our model on these cases in a separate evaluation by reporting whether residual tumour was predicted (false positive) and reporting the corresponding volume of falsely predicted tumour.

## 3 RESULTS

Example segmentations generated by the deep learning models trained on the different input modalities of the MC-RC dataset are shown in Figure 3. The selected example cases have DSCs close to the median DSCs achieved on the respective testing sets. The models correctly predict most of the tumour volume and deviate from the ground truth only in regions of low image contrast between tumour and surrounding tissues. Especially on T2w images, tumour boundaries are less pronounced and can be ambiguous even to human annotators as seen in the jagged through-plane contour lines of the ground truth in Figure 3b. Notably, the models avoid inconsistencies between adjacent slices in all spatial directions and render smoother tumour boundaries. Reduced performance on T2w images is consistent with the measured inter- and intra-observer reliability which was significantly higher for ceT1w images. The average DSC between two annotators was 88.1±3.4% (minimum: 87.5±4.3%, maximum: 89.1±3.4%) when the segmentation was performed on ceT1w images and 84.5±7.8% (minimum: 82.5±13.9%, maximum: 85.8±7.9%) when performed on T2w. Similarly, intra-observer reliability was higher for ceT1w (87.8±4.4%) than for T2w (84.4±11.5%).

**Figure 3.**
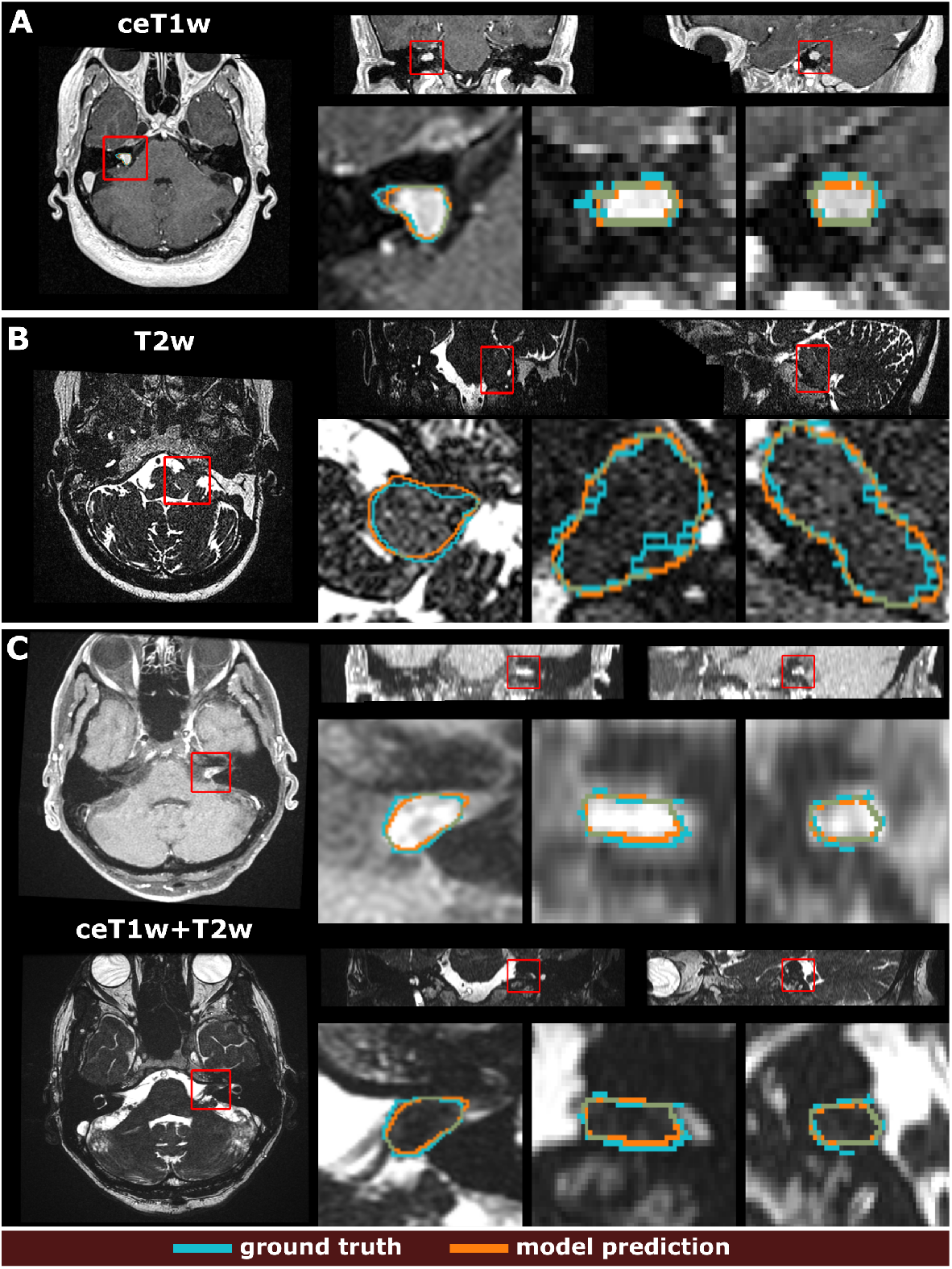
Example segmentation results generated with the 3 models trained on the multi-centre clinical (MC-RC) training sets. Predictions in **(A)** were obtained with the ceT1w model, predictions in **(B)** with the T2w model and predictions in **(C)** with the ceT1w+T2w model. Each example shows axial, coronal, and sagittal views of the full MRI image and magnified images of the tumour region. The magnified region is indicated by a red bounding box. Dice similarity coefficients (DSC) were 86.3%, 89.0%, and 87.1% respectively, which is close to the median DSC achieved by each model on the MC-RC testing set.

**Figure 4.**
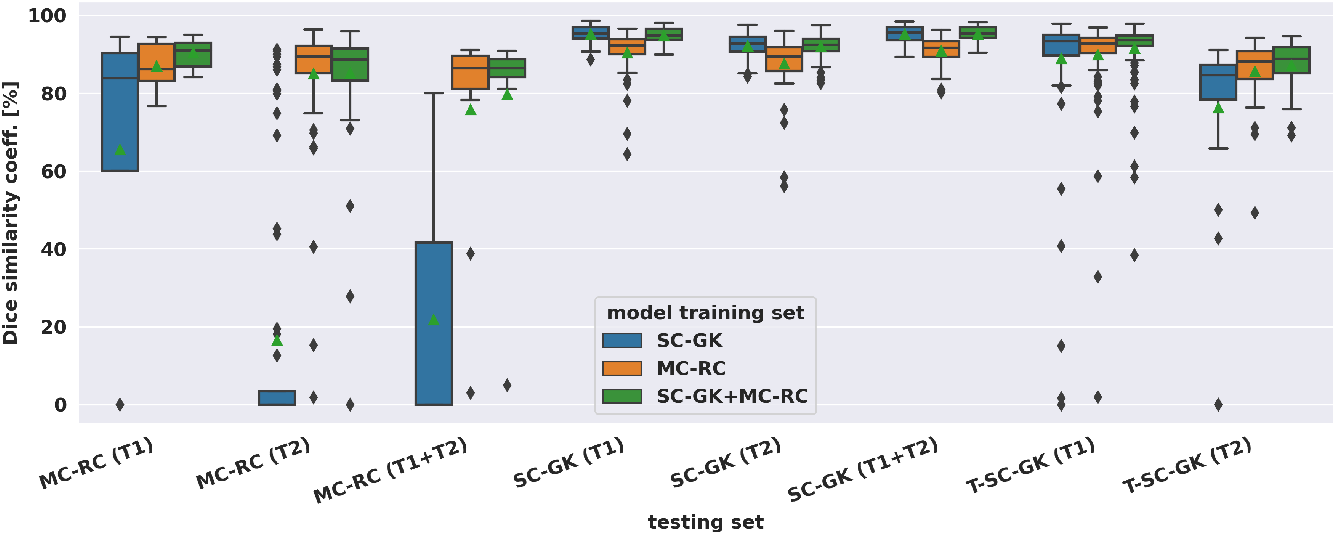
Dice similarity coefficients achieved by deep learning models on multi-centre clinical (MC-RC) and single-centre Gamma Knife (SC-GK) datasets. The x-axis indicates on which testing set the models were evaluated. The centre vertical line indicates the median and the green triangle indicates the mean. The boxes extend from the lower quartile *Q*_1_ to the upper quartile *Q*_3_, the whiskers extend from *Q*_1_ − 1.5(*Q*_3_ − *Q*_1_) to *Q*_3_ + 1.5(*Q*_3_ − *Q*_1_). Data beyond the whiskers are considered outliers and shown as black diamonds.

Mean DSCs achieved by all models on MC-RC, SC-GK, and T-SC-GK testing sets are presented in Table 1. Models which were trained on our MC-RC training sets performed well on all testing sets. The model performance in terms of average DSCs was comparable to that achieved by human annotators in our inter- and intra-observer experiments. In contrast, the models trained on the SC-GK training sets performed well only on the SC-GK and T-SC-GK testing sets but poorly on the MC-RC testing sets. This highlights that the variability of the MC-RC training set is key to obtaining robust segmentation results in a clinical setting. The combined model (SC-GK+MC-RC) performs best on most testing sets and results in the smallest number of failures. Its capability for generalization, can further be assessed on the independent T-SC-GK testing sets on which it outperformed the other models.

The spread of DSCs is shown in the box and whisker plots of Figure 4. Performance on ceT1w images was generally slightly better than on T2w. The MC-RC ceT1w+T2w model with combined input modalities performed worse on the corresponding MC-RC testing set than the MC-RC models for separate input modalities. This is due to the relatively small number of training cases available for the ceT1w+T2w model (n=48) and the presence of large cystic components in the testing set which the model failed to include in the segmentation as illustrated in Figure 5. Although the models trained on the MC-RC training sets generated some outliers there was always a partial overlap between ground truth and segmentation (DSC *>* 0) so that no tumour was missed completely. Failure modes of each MC-RC model with examples of the worst cases are addressed in the next section. In contrast, the number of outliers and complete misses (DSC=0) by the SC-GK models on the other testing sets (MC-RC and T-SC-GK) was significantly higher. Median DSCs and other commonly reported metrics for segmentation tasks are reported in Table 2.

**Table 2.**
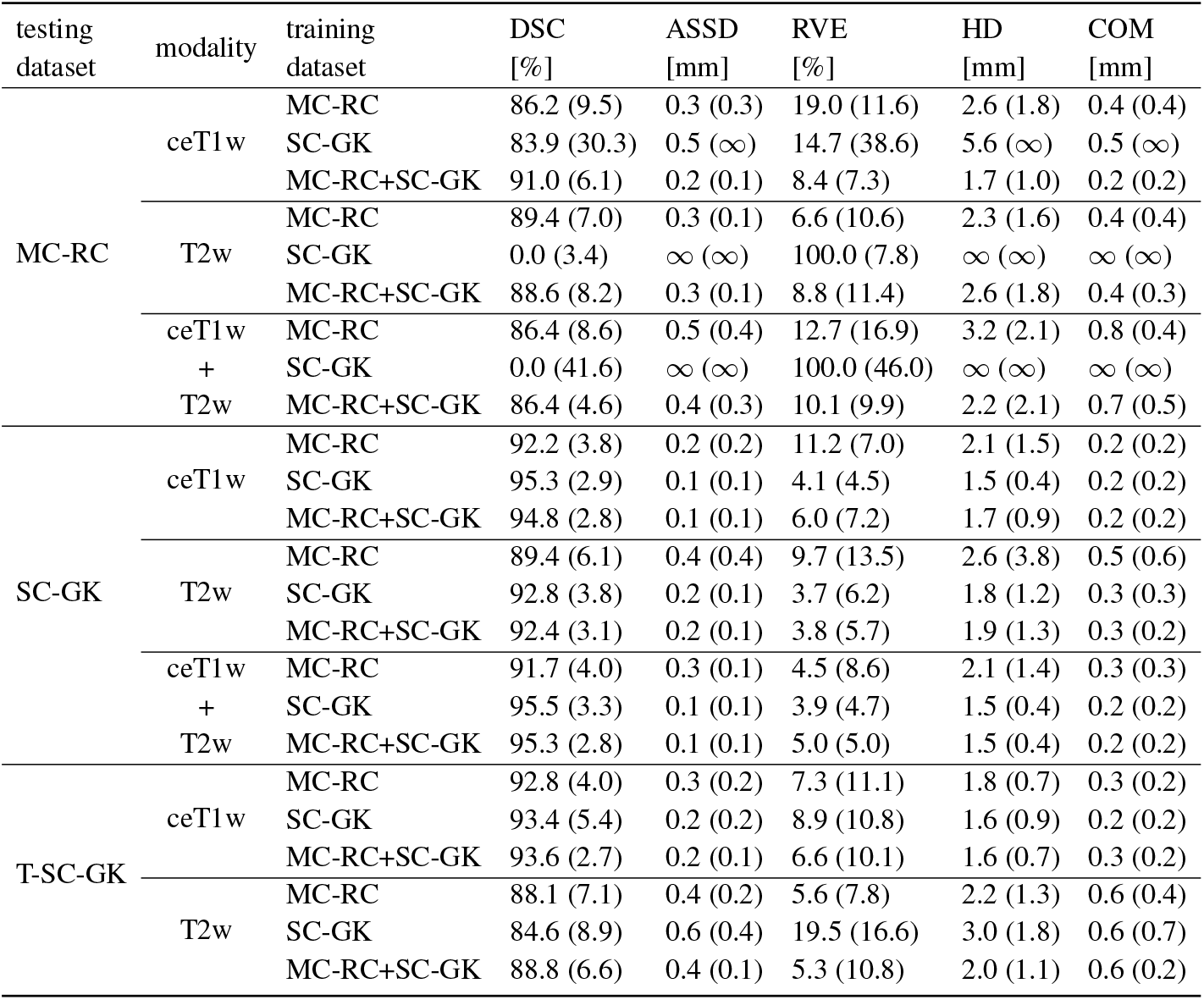
Additional commonly reported segmentation metrics (median and interquartile range). The values represent the median and interquartile ranges over all cases in the testing sets. The metrics are Dice similarity coefficient (DSC), average symmetric surface distance (ASSD), relative absolute volume error (RVE), undirected Hausdorff distance (HD), and distance between the centres of mass (COM). A ASSD, HD or COM value of ∞ (infinity) for a single case indicates that the model did not predict any VS in the image. A median value of ∞ shows that this occured in over half of the testing cases. In this case the metric is not defined.

| testing dataset | modality | training dataset | DSC [%] | ASSD [mm] | RVE [%] | HD [mm] | COM [mm] |
| --- | --- | --- | --- | --- | --- | --- | --- |
| MC-RC | ceT1w | MC-RC | 86.2 (9.5) | 0.3 (0.3) | 19.0 (11.6) | 2.6 (1.8) | 0.4 (0.4) |
| | | SC-GK | 83.9 (30.3) | 0.5 (∞) | 14.7 (38.6) | 5.6 (∞) | 0.5 (∞) |
|  |  | MC-RC+SC-GK | 91.0 (6.1) | 0.2 (0.1) | 8.4 (7.3) | 1.7 (1.0) | 0.2 (0.2) |
|  | T2w | MC-RC | 89.4 (7.0) | 0.3 (0.1) | 6.6 (10.6) | 2.3 (1.6) | 0.4 (0.4) |
| | | SC-GK | 0.0 (3.4) | ∞ (∞) | 100.0 (7.8) | ∞ (∞) | ∞ (∞) |
|  |  | MC-RC+SC-GK | 88.6 (8.2) | 0.3 (0.1) | 8.8 (11.4) | 2.6 (1.8) | 0.4 (0.3) |
|  | ceT1w + T2w | MC-RC | 86.4 (8.6) | 0.5 (0.4) | 12.7 (16.9) | 3.2 (2.1) | 0.8 (0.4) |
| | | SC-GK | 0.0 (41.6) | ∞ (∞) | 100.0 (46.0) | ∞ (∞) | ∞ (∞) |
|  |  | MC-RC+SC-GK | 86.4 (4.6) | 0.4 (0.3) | 10.1 (9.9) | 2.2 (2.1) | 0.7 (0.5) |
| SC-GK | ceT1w | MC-RC | 92.2 (3.8) | 0.2 (0.2) | 11.2 (7.0) | 2.1 (1.5) | 0.2 (0.2) |
|  |  | SC-GK | 95.3 (2.9) | 0.1 (0.1) | 4.1 (4.5) | 1.5 (0.4) | 0.2 (0.2) |
|  |  | MC-RC+SC-GK | 94.8 (2.8) | 0.1 (0.1) | 6.0 (7.2) | 1.7 (0.9) | 0.2 (0.2) |
|  | T2w | MC-RC | 89.4 (6.1) | 0.4 (0.4) | 9.7 (13.5) | 2.6 (3.8) | 0.5 (0.6) |
|  |  | SC-GK | 92.8 (3.8) | 0.2 (0.1) | 3.7 (6.2) | 1.8 (1.2) | 0.3 (0.3) |
|  |  | MC-RC+SC-GK | 92.4 (3.1) | 0.2 (0.1) | 3.8 (5.7) | 1.9 (1.3) | 0.3 (0.2) |
|  | ceT1w + T2w | MC-RC | 91.7 (4.0) | 0.3 (0.1) | 4.5 (8.6) | 2.1 (1.4) | 0.3 (0.3) |
|  |  | SC-GK | 95.5 (3.3) | 0.1 (0.1) | 3.9 (4.7) | 1.5 (0.4) | 0.2 (0.2) |
|  |  | MC-RC+SC-GK | 95.3 (2.8) | 0.1 (0.1) | 5.0 (5.0) | 1.5 (0.4) | 0.2 (0.2) |
| T-SC-GK | ceT1w | MC-RC | 92.8 (4.0) | 0.3 (0.2) | 7.3 (11.1) | 1.8 (0.7) | 0.3 (0.2) |
|  |  | SC-GK | 93.4 (5.4) | 0.2 (0.2) | 8.9 (10.8) | 1.6 (0.9) | 0.2 (0.2) |
|  |  | MC-RC+SC-GK | 93.6 (2.7) | 0.2 (0.1) | 6.6 (10.1) | 1.6 (0.7) | 0.3 (0.2) |
|  | T2w | MC-RC | 88.1 (7.1) | 0.4 (0.2) | 5.6 (7.8) | 2.2 (1.3) | 0.6 (0.4) |
|  |  | SC-GK | 84.6 (8.9) | 0.6 (0.4) | 19.5 (16.6) | 3.0 (1.8) | 0.6 (0.7) |
|  |  | MC-RC+SC-GK | 88.8 (6.6) | 0.4 (0.1) | 5.3 (10.8) | 2.0 (1.1) | 0.6 (0.2) |

**Figure 5.**
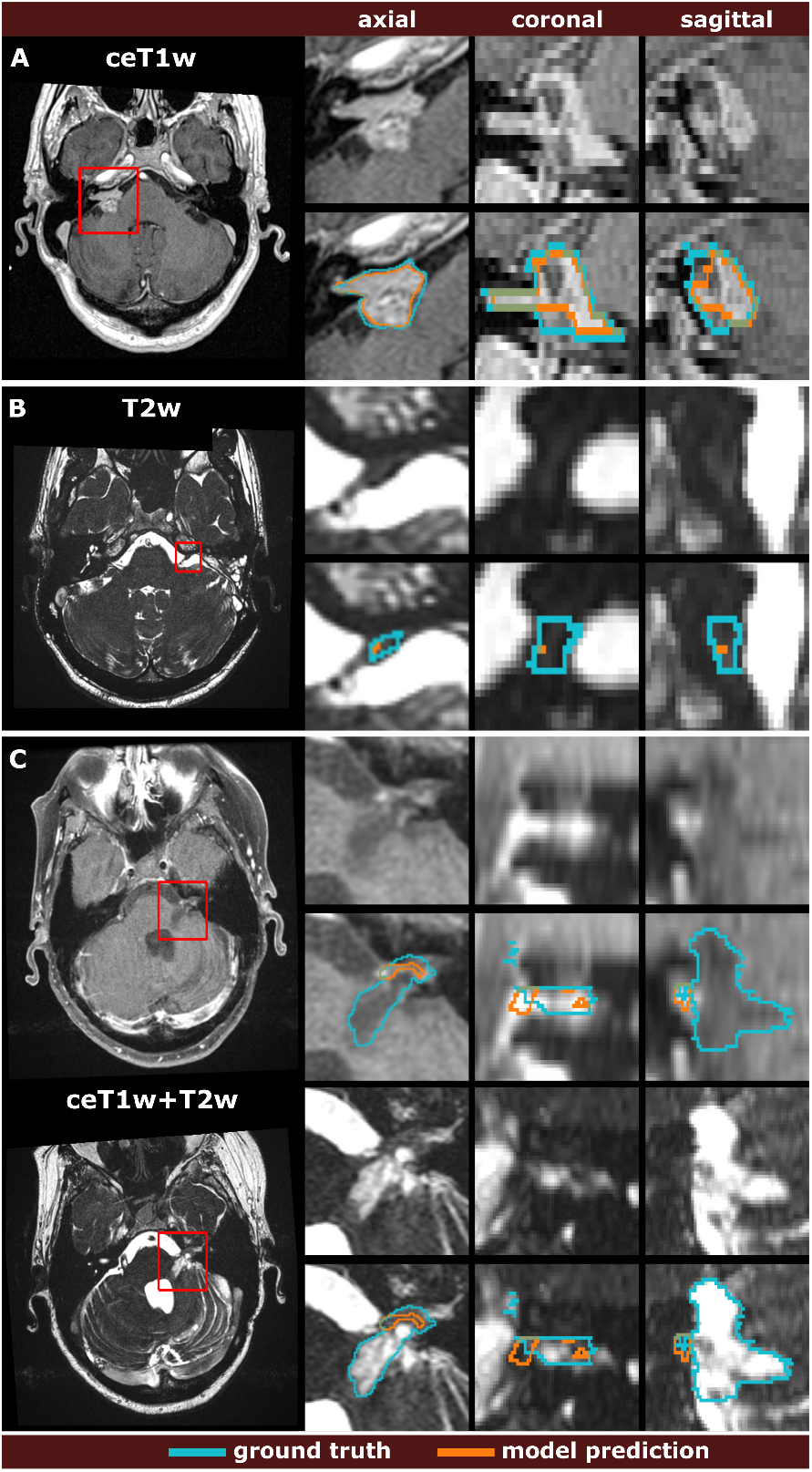
Comparison of the worst model predictions with the manual segmentation ground truth for each input modality. The models were trained on the multi-centre routine clinical (MC-RC) dataset using ceT1w images **(A)**, T2w images **(B)**, or their combination ceT1w+T2w **(C)**. Each example shows an axial slice of the full MRI image and magnified images of the tumour region. The magnified region is indicated by a red bounding box. Dice similarity coefficients were 73.6%, 1.9%, and 4.7% respectively.

Post-operative cases without residual tumour were assessed separately for the MC-RC models. For ceT1w input, all 7 cases were correctly labelled without residual tumour. For ceT1w+T2w input, a residual of 24 mm^3^ was predicted in 1 of 4 cases. Most false positives occurred for T2w input where 6 out of 9 predictions were in agreement with the ground truth, while the other 3 predictions suggested residuals of 1, 3, and 23 mm^3^, respectively.

## 4 DISCUSSION

### 4.1 Summary of contributions

For an automatic VS segmentation model to be useful in a routine clinical setting, accurate and reliable performance irrespective of acquisition parameters and tumour presentation is essential. In this work, we developed the first model for automatic VS segmentation whose application is not limited to MRI images from a specific scanner and acquisition protocol. Rather, by collecting a large multi-centre dataset and providing labour-intensive high-quality annotations, it was possible to train and evaluate a model that generalizes well under a wide range of settings and for all time points encountered in clinical routine, including initial diagnostic scans as well as scans of post-operative tumour residuals.

The generated automatic segmentations had average DSCs comparable to those of human annotators as measured by inter- and intra-observer experiments and performed robustly on independent datasets. Therefore, this work represents a key step toward the incorporation of automated segmentation algorithms in the clinical workflow and management of VS patients.

For example, based on the model segmentation, automatic surveillance of the patient’s tumour growth through longitudinal scans could be performed (Shapey et al., 2021a). Currently, in routine clinical practice, tumour size is usually assessed by determination of the maximum extrameatal linear tumour dimension, although several studies have shown that tumour volume is a more reliable and accurate metric to measure tumour growth (MacKeith et al., 2018; Walz et al., 2012; Tang et al., 2014; Roche et al., 2007). Using our deep learning model, the automatic calculation of tumour volume is a simple task. Moreover, the model could be used to generalize methods for automatic classification of VS according to the Koos scale which requires accurate tumour segmentations as an initial step (Kujawa et al., 2022; Koos et al., 1998). Finally, the model could be used as an initialization for interactive segmentation approaches (Wang et al., 2018b,a) or as input for subsequent models that further segment the VS into intra- and extrameatal components (Wijethilake et al., 2022).

A study similar to ours was published recently. Neve et al. (2022) employ a private multi-centre dataset of ceT1w and T2w images for model training and evaluation. In contrast to our study, no longitudinal imaging data but rather a single time point per patient were considered. Furthermore, all post-operative

images were excluded from the model training and analysis. Although the detection of residual tumour tissue is more difficult it is essential for post-operative surveillance and detection of tumour recurrence. A comparison of segmentation accuracy in terms of DSC is presented in Table 3.

**Table 3.** Comparison with state-of-the-art results in terms of mean Dice similarity coefficients and standard deviation. Prior results are based on data extracted from treatment plans for Gamma Knife stereotactic radiosurgery. Another study employs a multi-centre dataset with pre-operative images. In each referenced publication different training and testing datasets were used.

| reference | training set | testing set | DSC [%] |  |  |
| --- | --- | --- | --- | --- | --- |
|  |  |  | ceT1w | T2w | ceT1w+T2w |
| Shapey et al. (2019) | Gamma Knife | Gamma Knife | $93.4 \pm 4.0$ | $88.3 \pm 3.9$ | $93.7 \pm 2.8$ |
| Shapey et al. (2021a) | Gamma Knife | Gamma Knife | $94.5 \pm 2.2$ | $90.7 \pm 3.6$ | - |
| Wang et al. (2019) | Gamma Knife | Gamma Knife | - | $87.3 \pm 4.9$ | - |
| Lee et al. (2021) | Gamma Knife | Gamma Knife | - | - | $90 \pm 5$ |
| Neve et al. (2022) | Multi-Centre<br>(pre-operative) | Multi-Centre<br>(pre-operative) | $92 \pm 5$ | $87 \pm 6$ | - |
| This work | SC-GK | SC-GK | $95.2 \pm 2.2$ | $92.2 \pm 3.4$ | $95.2 \pm 2.0$ |
| This work | MC-RC | MC-RC | $86.9 \pm 5.9$ | $85.0 \pm 15.6$ | $75.8 \pm 27.0$ |

### 4.2 Comparison with state-of-the-art

Like the model presented here, state-of-the-art methods for VS segmentation employ CNNs based on a U-Net architecture. The referenced average DSCs as well as our DCSs listed in Table 3 were obtained on hold-out testing sets that were drawn from the same datasets as the training sets. Within the margin of error, our results obtained with the SC-GK dataset are comparable to or outperform previously reported results, confirming the validity of our model training approach. On the MC-RC dataset, DSCs are decreased as a result of the increased variability of the MC-RC testing sets compared to the more homogenous datasets used in the referenced studies. Especially the presence of post-operative cases with residual tumours increases the complexity of the segmentation task. This interpretation is supported by the reduced inter-observer reliability (DSC= 88.1 ± 3.4% on ceT1w images and 84.5 ± 7.8% on T2w images) compared to the inter-observer reliability reported on the SC-GK dataset (DSC=93.82 ± 3.08% based on ceT1w and T2w images). For ceT1w+T2w input, another contributing factor is the relatively small number of available training cases.

### 4.3 Worst cases

While the MC-RC models fail in only a small number of cases the analysis of the corresponding images and faulty predictions can highlight potential model weaknesses with regards to specific acquisition settings and tumour presentations. Figure 5 shows the worst model prediction for each of the three inputs. The ceT1w case inFigure 5a and T2w case in Figure 5b are post-operative scans after surgery with translabyrinthine approach. The model prediction for the ceT1w image has an acceptable DSC of 73.6% since it captures parts of the tumour with high contrast agent uptake but misses lower contrast regions in inferior slices. The T2w case contains a small tumour residual which is almost entirely missed by the model (DSC=1.9%). Due to the low contrast with adjacent tissue this tumour residual is particularly difficult to segment. For this case, a human annotator would require a contrast-enhanced scan to confirm the boundaries of the residual. The case shown in Figure 5c is a post-operative scan after surgery with retrosigmoid approach with a large cystic component. While parts of the solid tumour residuals are accurately delineated, the cystic component is missed by the model prediction (DSC=4.7%). It is likely that the small training set available for ceT1w+T2w input did not contain a sufficiently large number of cystic tumours to train the model with respect to their inclusion.

### 4.4 Post-operative cases without residual tumour

Cases without residual tumour are edge cases that can be particularly challenging for segmentation algorithms. While the MC-RC models correctly predicted no residual tumour in 16 out of 20 cases, small residuals (*<*30 mm^3^) were predicted for the remaining cases. While the ceT1w model was reliable, the T2w model led to false positive predictions in 3 of 9 cases. This difference in robustness is expected because residuals are typically hyper-intense on ceT1w images while they are difficult to discern in images without contrast enhancement.

A frequently applied post-processing strategy for segmentation models is to remove segments below a fixed volume threshold (Antonelli et al., 2022). A reasonable volume threshold can be based on an assumed detection limit for VS in routine clinical MRI of 2 mm, which corresponds to a cubic volume of 8 mm^3^. In comparison, the smallest tumour residual contained in the MC-RC dataset was 30 mm^3^. Application of this post-processing strategy improved the number of correct predictions to 18 out of 20 cases without significantly affecting the results presented in Table 1 and Table 2.

### 4.5 Limitations and future work

While the dataset curated and annotated in this work is the first large multi-centre dataset for VS segmentation made publicly available, sample sizes for ceT1w and ceT1w+T2w images were small compared to T2w. This is because the slice thickness of the majority of ceT1w images in routine clinical dataset exceeded the threshold of the inclusion criterion (3.9 mm). Detection and manual segmentation of small tumours on these low-resolution images is difficult and generally not sufficiently accurate for volumetric measurements. We expect that segmentation performance can be improved by increasing the number of ceT1w images in the dataset.

Furthermore, since this study focuses on sporadic unilateral VS, bilateral tumours in patients with the hereditary condition NF2 were excluded. Due to the simultaneous presence of multiple schwannomas and meningiomas, the segmentation task is disproportionately more difficult. In the future, we plan to integrate NF2 cases into the model development.

These models can be applied for the automatic generation of case reports for multidisciplinary team meetings (MDM) (Wijethilake et al., 2023). The reports in their work include multiple automatically generated views of the tumour and the model segmentation and frequently reported tumour measures, such as volume and extrameatal dimensions. It will be interesting to assess how the reports might facilitate, on the one hand, MDM preparation, and on the other hand, the treatment decision process during the meeting itself.

In conclusion, we developed a model for automatic VS segmentation for diverse clinical images acquired at different medical centres with a wide range of scan protocols and parameters. The application of this model has the potential to monitor tumour size, post-operative residuals, and recurrence more accurately and efficiently, thereby facilitating the VS surveillance and management of patients.

## Data Availability

The MC-RC dataset and all trained deep learning models were made available online.

https://doi.org/10.5281/zenodo.10363647

## CONFLICT OF INTEREST STATEMENT

SO, TV, and JS are shareholders and co-founders of Hypervision Surgical Ltd.

The authors otherwise declare that the research was conducted in the absence of any commercial or financial relationships that could be construed as a potential conflict of interest.

## FUNDING

This work was supported by Wellcome Trust (203145Z/16/Z, 203148/Z/16/Z, WT106882), EPSRC (NS/A000050/1, NS/A000049/1) and MRC (MC/PC/180520) funding. Additional funding was provided by Medtronic. TV is also supported by a Medtronic/Royal Academy of Engineering Research Chair (RCSRF1819/7/34). For the purpose of open access, the authors have applied a CC BY public copyright licence to any Author Accepted Manuscript version arising from this submission.

## ACKNOWLEDGMENTS

The authors would like to thank Dr Andrew Worth and Gregory Millington for their contributions to the generation of the segmentation ground truth.

## DATA AVAILABILITY STATEMENT

The datasets used in this study are available here:

- Multi-Center routine clinical (MC-RC): https://doi.org/10.7937/HRZH-2N82
- Single-Center Gamma Knife (SC-GK): https://doi.org/10.7937/TCIA.9YTJ-5Q73
- Tilburg Single-Center Gamma Knife (T-SC-GK): https://www.synapse.org/#!Synapse:syn51236108

The pre-trained models are available here: https://doi.org/10.5281/zenodo.10363647

## Notes

### Competing Interest Statement

Funding was provided by Medtronic. SO, TV, and JS are shareholders and co-founders of Hypervision Surgical Ltd.

### Author Declarations

This study was approved by the NHS Health Research Authority and Research Ethics Committee (18/LO/0532).

### Summary of Updates

The original MC-RC dataset was revised by further refining the segmentations and excluding additional images. All experiments were repeated based on the refined dataset. Additional models based on a combined training set and further validation experiments based public and private datasets were added.

